# A framework for human-artificial intelligence co-learning for disease activity labeling using electronic health records

**DOI:** 10.64898/2026.07.16.26358271

**Authors:** Zongxin Yang, Yuming Zhang, Zoe Love, Abisayo Animashaun, Katherine Zhong, Gregory McDermott, Tianxi Cai, Katherine P. Liao

## Abstract

**Objective:** To develop and evaluate a framework for human-AI interaction. This approach, SHARE (Synergistic Human-Agent REasoning system) was designed to support scalable phenotyping of complex outcomes accurately, robustly and reproducibly from real-world electronic health record (EHR) data to support real-world evidence (RWE) generation.

**Methods and Analysis:** Using rheumatoid arthritis (RA) disease activity as the use-case, we studied a multi-institutional EHR-based RA cohort of 3,167 patients. Expert reviewers and a disease activity agent labeled notes using the same review guideline. The agent combined embedding-based informative-note filtering, structured evidence extraction, and evidence-based integrated reasoning to assign disease activity categories with supporting evidence, rationale, confidence, and ambiguity flags. To support scalable deployment, we evaluated a budget-tiered configuration using GPT-5 Nano for high-volume evidence extraction, o4-mini for final reasoning, benchmarking against a GPT-5.4 high reasoning effort configuration applied at every step. Note-level discrepancies were adjudicated by reviewers into final co-produced labels that were used to refine labels and inform agent development. The main outcome measure was the mean absolute error (MAE) of the initial and final agent vs the final co-produced labels. The agreement between agent- and reviewer-flagged ambiguous notes, per-note cost and compute time across configurations were also tested.

**Results:** Expert reviewers labeled 626 notes from 273 patients; human-AI adjudication revised 127 (20%) of these initial labels and added 60 newly labeled notes, yielding a 686-note co-produced reference. Against this reference, the final agent’s accuracy improved from a mean absolute error of 0.406 to 0.291 with co-learning, and its ambiguity flag agreed with expert ambiguity designations with 92.1% accuracy. Applied across the cohort, the agent labeled 101,691 notes; the budget tiered configuration matched the accuracy of GPT-5.4 at high reasoning effort while reducing estimated cost by 69% and compute time by 70%.

**Conclusion:** Adopting a framework for human-AI co-learning, SHARE, improved the overall quality of gold-standard labels, identified ambiguous cases for further review, and supported accurate and standardized chart reviews of disease activity at a scale infeasible for manual review. SHARE’s resource efficiency provides a transferable approach to incorporate complex phenotypes in RWE studies.

**Key messages:** *What is already known on this topic:* Defining disease states from electronic health record (EHR) data is central to generating real-world evidence (RWE), but complex phenotypes require extensive review of narrative clinical notes that are difficult to standardize, audit, and scale. Out-of-the-box large language model (LLM) prompting can support review, but accurate annotation with face validity requires workflows that preserve supporting evidence, recognize uncertainty, while keeping the clinical experts in the adjudication loop.

*What this study adds:* We developed and evaluated the Synergistic Human-Agent REasoning system (SHARE), a multi-stage human-artificial intelligence (AI) co-learning framework in which clinical experts define phenotype guidelines and the agent identifies informative notes, extracts supporting evidence, assigns labels, and flags ambiguity for focused review. With rheumatoid arthritis disease activity as a use-case, adjudication improved and expanded the reference labels, while selective use of lower- and higher-cost models supported internally evaluated, resource-efficient scaling.

*How this study might affect research, practice or policy:* SHARE introduces a framework for human-AI workflows for research, shifting review of complex EHR phenotypes from broad manual abstraction towards a scalable, resource-efficient, targeted expert adjudication, with clinical experts defining the guidelines, overseeing local validation and the final interpretation.

## Introduction

Real-world electronic health record (EHR) data can complement traditional randomized controlled trials (RCTs) by studying populations that are larger and more diverse and/or by investigating outcomes that are rare or difficult to study in an RCT within a feasible time-frame^1,2^. For many chronic conditions, however, a major bottleneck for generating real-world evidence (RWE) for treatment effects is a lack of a standardized measure for key clinical outcomes, such as disease activity level, at-scale. In RCTs of treatment interventions, key study end points typically measure outcomes at multiple time points. Across conditions, disease activity is a composite score that incorporates exam findings, questionnaires, or laboratory data. In RCTs, the components of disease activity are directly measured in-person and calculated at study visits. In routine clinical care, information on disease activity is typically dispersed across different sections of unstructured narrative notes and laboratory test results, and clinical impressions, often recorded with variable terminology and clinical context.

This lack of structure creates problems with scalability and determining uncertainty. Human clinical experts can infer disease activity through chart review, however, such reviews are time-intensive, can be inconsistent, and are infeasible for large-scale longitudinal studies. Moreover, many notes do not support a single definitive label. Disease activity may be ambiguously documented, supported by conflicting evidence, or described indirectly through treatment decisions, symptoms, laboratory trends, or physician judgment. For complex phenotypes such as rheumatoid arthritis disease activity, accurate labeling therefore requires more than extracting isolated concepts from text; it requires identifying the relevant evidence, integrating heterogeneous clinical signals, applying disease-specific rules, and explicitly recognizing when the available evidence supports uncertainty rather than a definitive category.

Large language models (LLMs) offer a potential way to standardize and scale complex EHR review, but out-of-the-box prompting remains insufficient in accurately assigning a disease state for phenotypes that require nuanced clinical reasoning and recognition of uncertain cases^3^. In addition, applying high-capability LLMs indiscriminately to large EHR corpora can be prohibitively expensive and computationally inefficient, particularly when most clinical notes contain little or no information relevant to a specific disease activity assessment. Thus, a practical framework for LLM-assisted large-scale EHR reviews for real-world data (RWD) must address not only accuracy, but also ambiguity, domain expert oversight, and cost-aware deployment. Such a workflow should also preserve note-level supporting evidence so that its outputs can be audited, corrected, and refined during clinician adjudication.

To address these challenges, we propose the Synergistic Human-Agent REasoning System (SHARE): a human-AI co-learning framework leveraging the strengths of clinical experts and LLMs. This framework includes the development of a Disease Activity (DA) agent. SHARE deconstructs the disease activity labeling into clear evidence-grounded steps: identifying informative notes, extracting clinically relevant evidence, assigning disease activity levels, and flagging ambiguous cases that can be further followed up with expert review.

To use computational resources efficiently, SHARE uses cost-effective LLMs for different steps of the workflow, reserving more resource-intensive processing for cases that require deeper reasoning. When the agent and human outputs disagree, human experts review the cases through an iterative adjudication to refine the initial reference labels. The adjudicated cases were then used to improve the agent, while the DA agent can apply the learned abstraction workflow across large note corpora, enabling standardized chart review at a scale infeasible for manual review. The product is a system that supports human-AI/LLM collaboration enabling extraction of complex disease specific outcomes, preserving expert judgement where uncertainty is high while using cost-aware strategies to make large-scale deployment feasible. In this study, we use disease activity for rheumatoid arthritis (RA), a chronic inflammatory condition, as a use-case to illustrate how human-AI co-learning can support scalable, standardized, ambiguity-aware, and resource-efficient labeling of complex clinical outcomes from real-world EHR data. Rather than replacing expert review, SHARE turns agent outputs into reviewable work products and concentrates clinician effort on disagreements, missing evidence, and intrinsically ambiguous notes. We internally evaluated whether co-learning improved agreement with the co-produced reference, whether ambiguity flags supported focused expert review, and whether a budget-tiered configuration reduced cost and compute time while preserving accuracy.

## Materials and Methods

### The SHARE co-learning framework

SHARE was developed to serve as a general human-AI co-learning framework for generating reproducible, expert-quality labels from unstructured clinical text at-scale (Figure 1A,B). In SHARE, clinical experts first define the labeling task and develop a written chart-review guideline to standardize reviews among members of the chart-review team (standard for traditional studies with ≥1 reviewer). The chart-review guideline is then used by the LLM agent to produce for each note, a structured label together with the supporting evidence it extracted, a rationale, and a confidence score. When the evidence is conflicting or incomplete the agent generates an explicit ambiguity flag with an alternative label. Notes where the expert and the agent disagree are returned to the experts, who adjudicate using the agent’s evidence and rationale.

**Figure 1.**
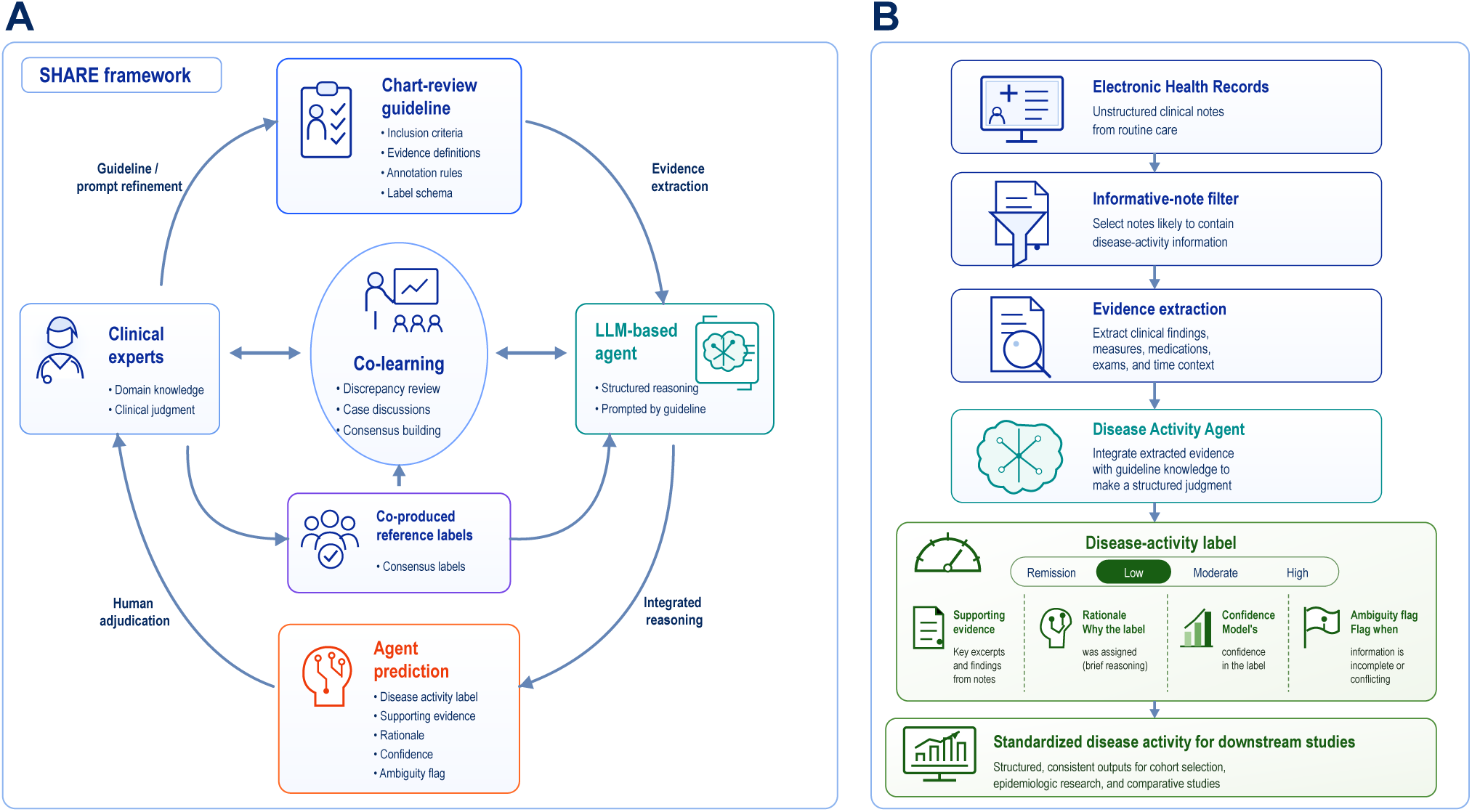
SHARE framework and SHARE-DA workflow. (A) Human-AI co-learning loop: clinical experts define chart-review guidelines; the LLM agent performs evidence extraction and integrated reasoning; and human adjudication informs guideline/prompt refinement and co-produced reference labels. (B) SHARE-DA applies this workflow to EHR notes through informative-note filtering, evidence extraction, and disease-activity reasoning, returning structured disease-activity labels (remission, low, moderate, high disease activity) with supporting evidence, rationale, confidence, and ambiguity flags for downstream studies.

The guideline and the agent’s prompts and rules are then refined. Iterating this loop yields two products: an adjudication-informed co-produced reference set and an internally evaluated agent configuration that can apply the learned abstraction to the entire note corpus. Because the framework is defined independently of any single disease, it is intended to transfer to other conditions and complex phenotypes.

### Study cohort and reference standard

We demonstrated SHARE on RA disease activity in the Mass General Brigham (MGB) multi-institutional EHR. The cohort mirrored a study comparing second-line therapies for RA of patients initiating a first tumor necrosis factor inhibitor (TNFi) on or after January 1, 2006 who subsequently switched to another biologic or targeted synthetic disease-modifying antirheumatic drug (b/tsDMARD), followed through August 2023 (Figure 2).

**Figure 2.**
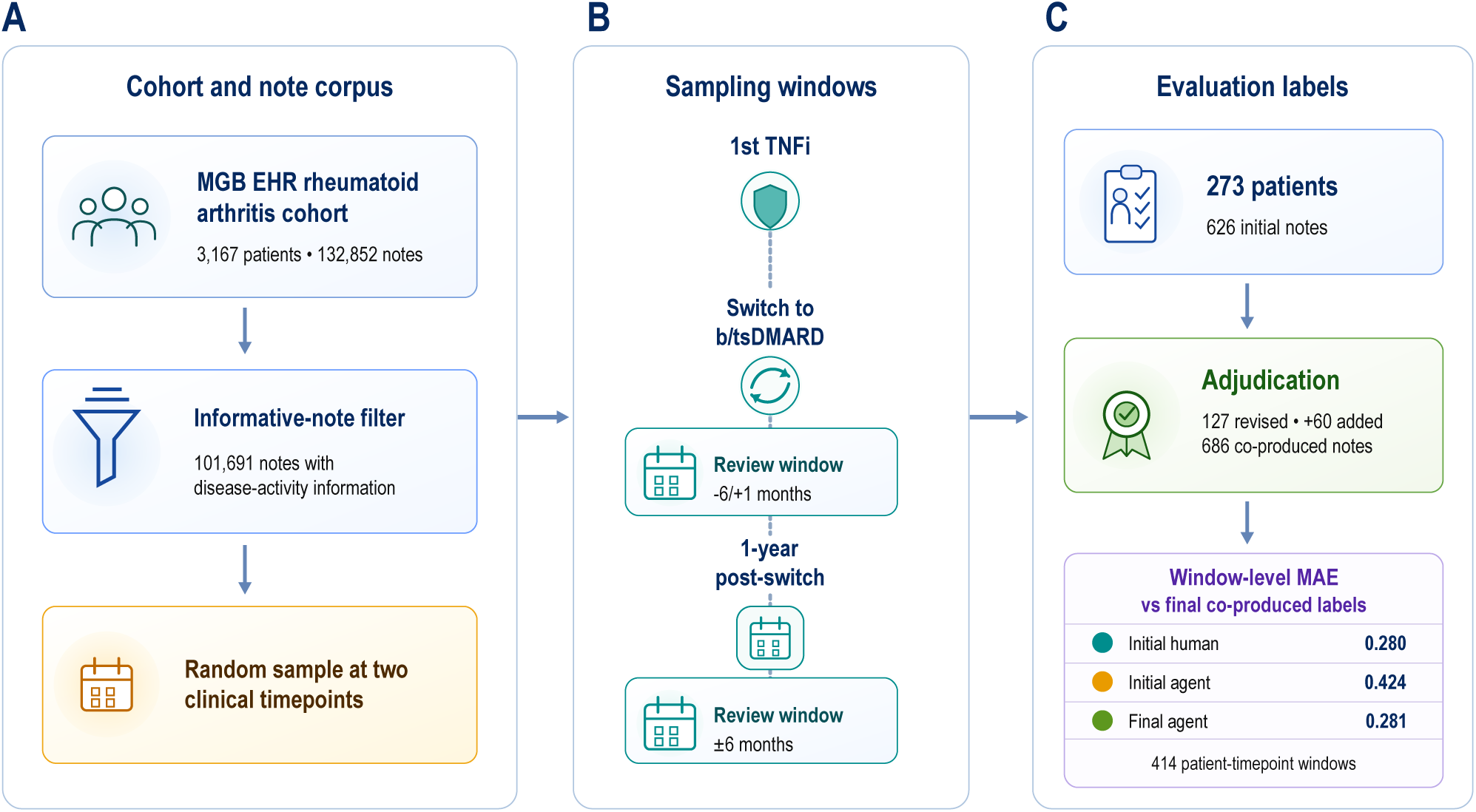
Overview of cohort sampling and comparison groups to evaluate SHARE-DA relative to human expert reviewers.

The clinical expert generated guideline assigned each note one of four ordinal disease-activity categories: remission, low, moderate, or high, approximating the disease activity score 28 using C-reactive protein (DAS28-CRP), a validated RA disease activity instrument used in clinical trials^4^. The guidelines additionally permitted clinical reasoning in addition to the calculated DAS28-CRP, for example, escalating the category when treatment was escalated.

To build the reference standard, we randomly sampled subjects at two clinically anchored timepoints: (1) at the time of the switch from TNFi to another b/tsDMARD, and (2) one year after the switch. The expert reviewers assigned a category to the most relevant notes nearest each timepoint, providing the initial human reference. If the reviewers were uncertain of the disease activity category due to conflicting or incomplete information, they flagged a note as ambiguous, recording both a primary and a secondary category.

### The SHARE-DA agent

The SHARE-DA agent is a chained module with three steps: (1) *Informative-note filtering.* In this step, an embedding-based filter screens out EHR notes which contain no disease-activity information; this comprises the vast majority of notes. A library of sentences relevant to disease activity was generated from passages identified by GPT-5 as containing core information relevant to RA disease-activity assessment in development notes; duplicate passages were removed to form the positive evidence library. Each candidate note was then segmented into overlapping 256-token chunks with 64-token overlap using the embedding-model tokenizer and compared with this library by cosine similarity. The maximum similarity across chunks defines the note’s informativeness score, thresholded at a value fixed on a held-out split of the GPT-5- labeled development set. The fixed informative-note filter was then evaluated on a separate clinician-labeled validation set before being applied to the RA corpus. (2) *Evidence extraction*. From each informative note the agent extracts the evidence required to assess disease activity, e.g., tender and swollen joint counts, any documented disease activity score (DAS), CRP, the physician global assessment, and treatment changes, each linked to a supporting excerpt. (3) *Evidence-based integrated reasoning*. In this step, the agent integrates all relevant evidence to assign a primary category, applying the prespecified expert rules alongside the calculated DAS28-CRP, and returns an ambiguity flag with a secondary category when the evidence is conflicting or incomplete. Every output is accompanied by supporting evidence, a rationale, and a confidence score for clinical expert review. More details about the implementation of SHARE-DA agent can be found in Supplementary Methods 1, the full output schema in Supplementary Table 1, and the prompt templates in Supplementary Methods 2.

### Cost-aware design

A central design goal was to reach expert-level accuracy at a cost and speed compatible with large-scale deployment for a research team, rather than to maximize accuracy regardless of resources. The proposed configuration reflects the cost-aware design which includes an embedding-based informative-note filter that screens out notes carrying no disease-activity signal, paired with a tiered model strategy that uses a lightweight model (GPT-5 Nano) for high-volume evidence extraction and a budget reasoning model (o4-mini, medium reasoning effort) for the final inference. We adopted this configuration specifically to make accurate labeling affordable at-scale.

To confirm that this budget-driven default does not come at the cost of accuracy, we benchmarked it against three alternatives, defined in Supplementary Methods 3. The first benchmark was a naive single-prompt baseline: the note and guideline are placed in one prompt. The model is then asked directly for a category, with no structured extraction. The second benchmark was a single-call version of the agent, using the same structured extraction-and-reasoning, completed in one model call. The third was a full multi-step agent run on a more powerful backbone, using GPT-5.4 at high reasoning effort, a more expensive model than o4-mini, applied at every step of the pipeline, representing what one might do without budget constraints. Accuracy, quantified by the mean absolute error (MAE) against the final human-AI co-produced labels, incorporating each method’s primary and secondary predictions, is reported alongside per-note cost and compute time (see Figure 3 and Supplementary Table 2 for more details).

**Figure 3.**
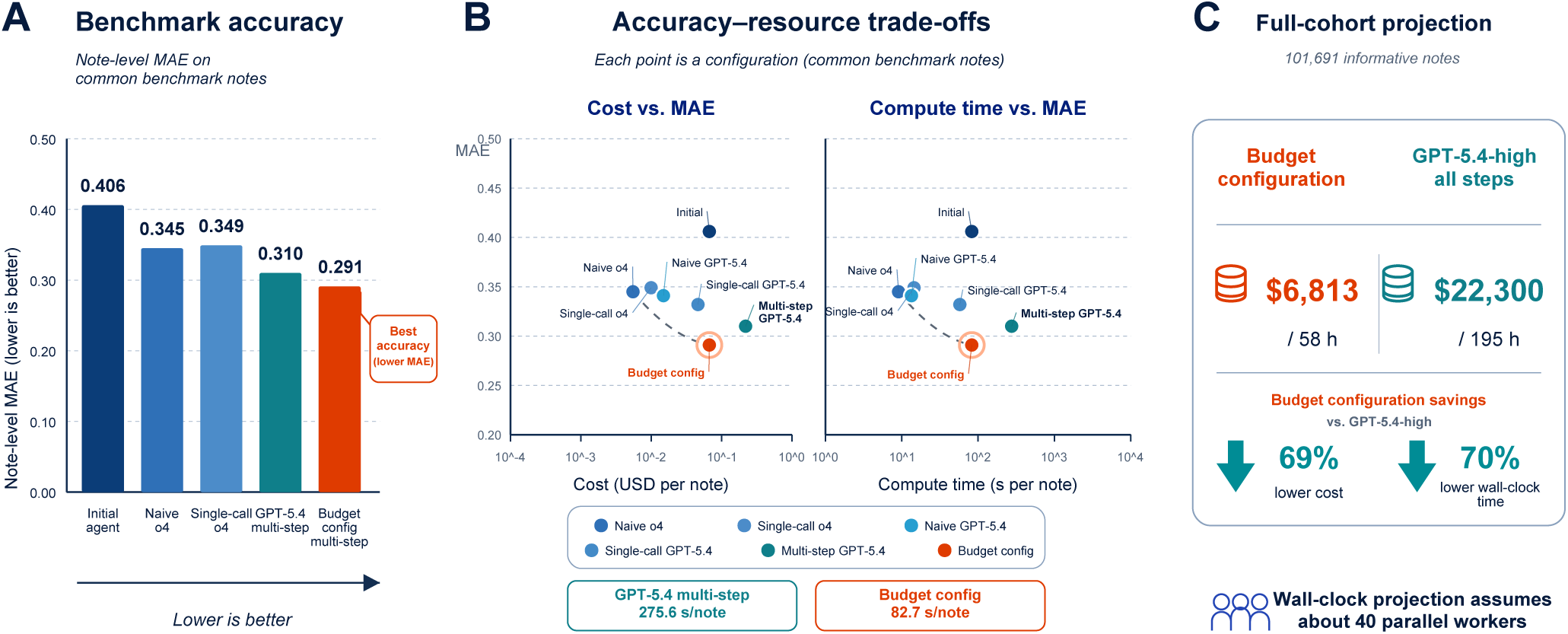
Benchmark accuracy, resource trade-offs, and full-cohort projection. (A) Note- level mean absolute error (MAE; lower is better) on the common benchmark set for the initial agent and benchmark/final configurations; “Budget config multi-step” represents the final agent used in the study. (B) Plots of MAE versus per-note cost and compute time; each point is a configuration, and the final budget configuration is highlighted. (C) Projected processing of 101,691 informative notes comparing the final budget configuration with GPT-5.4-high at all steps; wall-clock time assumes about 40 parallel workers and represents projected batch processing, not deployment.

### Human-AI adjudication and co-learning

Disease-activity category labels assigned from the initial human review and from the agent were compared note by note, and discrepancies were returned to the clinical experts for adjudication. For each discrepant note, reviewers examined the original note together with the agent’s predicted category, extracted evidence, rationale, confidence, and ambiguity flag, and recorded a final co-produced label. Adjudication findings were used to inform cautious refinement of the chart-review guidance and agent behavior during development.

### Outcomes and statistical analysis

The primary analysis was conducted comparing the mean absolute error (MAE) of the initial human annotation, the initial agent, and the final agent compared to the final co-produced labels. The disease activity category label provided by the human and agent were mapped to an ordinal score (remission = 1, low = 2, moderate = 3, high = 4). For ambiguous labels, the score was the confidence-weighted average of the primary and secondary scores (weighted by the reviewers’ assigned probabilities for the reference or by the agent’s confidence for its predictions). Accuracy is reported at two levels: (1) Note-level MAE (the level at which the reference was adjudicated) underlies the co-learning improvement and the comparisons on adjudicated and ambiguous notes; (2) window-level MAE summarizes disease activity over a recency-weighted time window around each clinical timepoint. This window-level assessment better reflects the accuracy in a real-world setting where humans and the agent are reviewing charts in a specific clinical time window of interest rather than specific notes.

For the primary analysis, we compared the MAE of the initial human chart review, the initial agent, and the final agent against the final co-produced labels; MAE=0 represents exact agreement. We additionally compared the MAE against the benchmark agent configurations (defined above under “Cost-aware design”). To understand the potential trade-off between cost and accuracy, the cost per note was plotted against the MAE for the different agent architecture and LLM backbone. Similarly for resource use, the compute-time was plotted against MAE.

We additionally aggregated note-level scores into recency-weighted patient-timepoint windows and recomputed MAE at this window level (see Supplementary Table 3 for more details on methods). Agreement on ambiguity was assessed by comparing the agent’s and reviewers’ ambiguity flags (accuracy, sensitivity, and specificity).

Prompt templates, implementation details, and versioning records were maintained in an access-controlled version-controlled repository.

### Patient and Public Involvement

Patients and the public were not directly involved in the study design, conduct, reporting, or dissemination plans.

## Results

### Study population

We studied data from 3,167 RA patients and a total of 132,852 notes. The mean age of the cohort was 61.4 years, 81.2% female and 70% were seropositive (rheumatoid factor or antibodies to cyclic citrullinated peptide positive). The informative note filter identified 101,691 notes with information on disease activity. The reviewers reviewed the records of 273 patients and provided disease activity labels for 686 notes in total.

### Human-AI co-learning refined both the reference and the agent

Disagreement between the initial agent and initial human labels was observed in 127 (20%) of the 626 initial human labels. Adjudication resulted in 87 definitive corrections to the initial human labels, almost all between adjacent categories, and 40 reclassified as ambiguous. The most common reason (78%) for updating an initial human disease activity label was a lack of standardization across reviewers in assessing joint involvement. An additional 60 notes from the same patients were missed by the human reviewers and were subsequently assigned a label leading to a total of 686 notes with labels.

Comparing against the final co-produced labels for disease activity, the final agent reached a note-level MAE of 0.291, an improvement over the initial agent’s MAE of 0.406 (Table 2). Among the challenging cases requiring adjudication (Table 2), the final SHARE-DA agent had a lower MAE (0.352) than the initial human reviews (0.847) when compared to the final co-produced labels.

### A budget-tier agent matched stronger models at a fraction of the cost

Across the common benchmark set, the final SHARE-DA agent achieved the lowest MAE (0.291), below the initial agent (0.406), o4-mini naive and single-call variants (0.345 and 0.349), and GPT-5.4-high multi-step (0.310; Figure 3A).

Accuracy-resource plots showed the final budget configuration with lower MAE, per-note cost, and compute time than the GPT-5.4-high multi-step configuration; within each backbone, multi-step reasoning improved accuracy relative to naive and single-call variants (Figure 3B).

Projected to the full cohort of 101,691 informative notes, the final budget configuration would cost approximately $6,813 and 58 hours across about 40 parallel workers, compared with $22,300 and 195 hours for GPT-5.4-high at every step — a 69% reduction in cost and 70% reduction in wall-clock time (Figure 3C; derived from the per-note costs and times in Supplementary Table 2).

### Ambiguity detection followed by focused expert review

Among 265 notes with both human and agent ambiguity assessments, human reviewers flagged 50 notes as ambiguous and the agent flagged 55. Both flagged 42 notes; the agent missed 8 reviewer-flagged ambiguous notes and flagged 13 additional notes not marked ambiguous by reviewers. This corresponded to 92.1% overall agreement, sensitivity of 0.840, and specificity of 0.940 (Table 1). Thus, agent-based triage would route 55 of 265 notes for focused review while capturing 42 of 50 expert- flagged ambiguous notes.

**Table 1:** Comparison of notes flagged as ambiguous by SHARE-DA compared to notes considered ambiguous by human reviewers.

|  |  |  |  |  |
| --- | --- | --- | --- | --- |
|  |  | Human annotation |  | Total |
|  |  | Ambiguous | Not ambiguous |  |
| Final SHARE-DA | Ambiguous | 42 (15.85%) | 13 (4.91%) | 55 (20.75%) |
|  | Not ambiguous | 8 (3.02%) | 202 (76.23%) | 210 (79.25%) |
| Total, # notes (%) |  | 50 (18.87%) | 215 (81.13%) | 265 (100%) |

Among the 50 reviewer-flagged ambiguous notes are the notes reclassified from a definite to an ambiguous category during adjudication (Results).

**Table 2:** Agent note-level accuracy before and after co-learning (mean absolute error vs the co-produced reference)

|  |  |  |
| --- | --- | --- |
| Note set | Approach | MAE |
| All evaluated notes | Initial SHARE-DA | 0.406 |
|  | Final SHARE-DA | 0.291 |
| Adjudicated subset (n = 120) | Human initial review | 0.847 |
|  | Initial SHARE-DA | 0.628 |
|  | Final SHARE-DA | 0.352 |
| *Initial human review is shown only for the adjudicated subset; because the co-produced reference was formed by revising the initial human labels, on the unrevised majority of notes the initial label equals the reference by construction, so an all-notes MAE would be driven mechanically toward zero (see Supplementary Table 3). All evaluated notes are the reference notes labeled by every configuration (Supplementary Table 2).* |  |  |

The MAE on the human-ambiguous notes was 0.45, about 50% higher than the overall note-level MAE of 0.291 (Table 2). This demonstrates that ambiguous notes explain a large part of the disagreement between the human and agent labels. The ability of the agent to flag uncertain cases for review with good accuracy allows it to select specific notes for focused expert review in place of a random large-scale review to achieve the same high quality disease activity designations.

### Accuracy against expert review at the window level

When disease activity was summarized over a time period around a clinical date rather than a single note, across 414 recency-weighted patient-timepoint windows, the final agent was comparable to expert review (MAE 0.281 vs 0.280; Figure 2).

## Discussion

In summary, SHARE provides a framework and process for investigators to interact and co-learn with LLM agents as part of existing clinical research workflows. The product of this collaboration includes accurate and standardized longitudinal information on complex disease state outcomes obtained efficiently at-scale, meeting a large unmet need for studies comparing treatment effects with RWD. We applied the framework to disease activity, largely available in the EHR narrative notes and an important component for defining treatment response across chronic conditions ^5,6^. SHARE is bidirectional: clinicians refine the disease activity agent through evidence-assisted adjudication and rule refinement. The agent supports the workflow by identifying information missed during initial review and applies the updated logic to a large corpus of notes that would be infeasible to review again. SHARE additionally had the capability to identify ambiguous cases, as the number of ambiguous cases is important for downstream interpretation of data. Lastly, we demonstrated that the framework can be optimized for resource efficiency while maintaining accuracy by reserving stronger LLMs for complex reasoning tasks, improving the accessibility of these technologies for research groups.

Clinical research with RWD has established methods for selecting appropriate cohorts, and gold standards for training algorithms to define phenotypes and outcomes^7,8^. While there is a growing body of evidence investigating LLMs for integration into clinical care, to our knowledge, limited frameworks exist for integrating LLMs into a clinical research workflow ^9–11^. Without SHARE, LLMs can be applied separately to assign disease activity per note, with prompts refined manually and iteratively in a process that can be challenging to track, evaluate, and scale. The SHARE framework includes a process that inserts interaction with LLMs as part of the workflow of a human research team.

The steps involving the initial agent mirrored what human chart reviewers do to achieve inter-rater reliability. Specifically, SHARE presented evidence supporting its disease activity classification to a reviewer to resolve cases where there was disagreement; if the agent was incorrect its logic could be updated to align with the human expert. Similarly, when two human reviewers disagree, the point for disagreement is discussed and the human logic for designating one disease activity category vs another is updated.

Importantly, the framework builds in an explicit process for internally evaluating and auditing LLM output^12^. Once the final configuration has been internally evaluated, SHARE can in effect serve as an additional reviewer-like tool, refined through human expert adjudication. We observed that the final agent can review thousands of notes in a standardized manner, more comprehensively than a human reviewer, with the same level of accuracy.

Recognizing uncertainty when considering a disease or disease state is an important part of clinical care and research. A thought process that occurs naturally in humans requires encoding and is an active area of study in AI^9,13–15^. Commercially available LLMs demonstrated good accuracy for assigning a final clinical diagnosis, but poor performance for generating differential diagnoses and reasoning in uncertain cases^16^. We demonstrated SHARE’s accuracy in flagging ambiguous cases, learning from the human labeling guidelines and labels. We observed that ambiguity was the main factor in the small discrepancies between the final agent disease activity labels and the final co-produced labels.

LLM-based systems that interact with expert reviewers can assist with clinical trial selection for common phenotypes such as heart failure, with initial input from clinical experts^17^. The SHARE framework adds to our existing knowledge by addressing needs in the study of complex phenotypes within a clinical research workflow, supporting iterative refinement which is needed even among human expert reviewers, and quantifying uncertainty. We demonstrate that these complex tasks can be accomplished in a resource-conserving manner^18^.

Study limitations include the use of one multi-institutional EHR dataset and demonstration of one type of disease activity as a concept. The co-produced reference was generated through unblinded adjudication with access to agent outputs, so this study does not provide independent blinded validation of the final agent. Future external evaluations should use frozen configurations and independently adjudicated reference labels. Future directions include transporting this framework and agent to independent healthcare systems and outcomes in other chronic conditions.

## Data Availability

Data available upon request and review and approval by the MGB Institutional Review Board.

## Contributors

Zongxin Yang conceived the study framework, developed the software and methods, performed the analyses, led the preparation and iterative refinement of the figures, and drafted and revised the manuscript. Yuming Zhang contributed to methodology, data curation, formal analysis and validation, and drafted and revised the manuscript. Zoe Love, Abisayo Animashaun, Katherine Zhong and Gregory McDermott contributed to clinician review and the human-AI co-learning process. Tianxi Cai and Katherine P. Liao contributed to study conceptualization and methodology, supervised the work, interpreted the results and critically revised the manuscript. All authors reviewed and approved the final manuscript. Katherine P. Liao is the guarantor.

## Ethics approval

This study was approved by the Mass General Brigham Institutional Review Board (protocol 2025P000263, “Artificial intelligence for phenotyping and enhancing clinical research methods with electronic health record data”) on September 30, 2025. The requirement for informed consent was waived.

## Use of artificial intelligence

OpenAI GPT-5.6 was used to assist with language editing and submission formatting. The authors reviewed and verified the resulting text and take full responsibility for the final content.

## Funding

This study was supported by the NIH R01AR080193, P30AR072577, K24AR085342

## Data Sharing Statement

Data available upon request and review and approval by the MGB Institutional Review Board.

## Conflict of Interest Disclosures

Dr Zhang reported receipt of grants from the Swiss National Science Foundation. Dr McDermott acted as site investigator for Boehringer Ingelheim clinical trial in areas unrelated to this study. Dr Liao consulted for UCB, Merck, BMS in areas unrelated to this study.

## Notes

### Author Declarations

The Mass General Brigham Institutional Review Board gave ethical approval for this work (protocol 2025P000263). The requirement for informed consent was waived.

