## Supplementary Information for "A framework for human-artificial intelligence co-learning for disease activity labeling using electronic health records"

**Supplementary Methods 1. SHARE-DA agent implementation**

SHARE-DA is implemented as a chained module; the complete per-note output schema is given in Supplementary Table 1. The three steps below extend the summary in the main text with the specific signals extracted and the reasoning pathways used.

**Informative-note filtering.** The informative-note filter was implemented as a separate retrieval-based classifier rather than a generative disease-activity call. Development labels were generated by GPT-5. Each note was passed to GPT-5 with a simple prompt asking whether the note contained core information relevant to rheumatoid arthritis disease-activity-score assessment. For notes labeled informative, GPT-5 also identified the supporting passage. Duplicate supporting passages were removed and indexed as the positive evidence library. Each candidate note is divided into overlapping 256-token chunks with 64-token overlap using the embedding-model tokenizer; each chunk and library passage is embedded with BAAI/bge-large-en-v1.5, L2-normalized, and compared with the library by inner product/cosine similarity, and the maximum cosine similarity across chunks is the note's informativeness score. The GPT-5-labeled development set contained 765 notes sampled from the same underlying RA-note source as the remaining corpus but not otherwise manually labeled for this study. Eighty percent of these notes (n = 612) were used to construct the positive evidence library, and the remaining 20% (n = 153) served as the held-out set for threshold selection by the Youden index on the receiver operating characteristic (ROC) curve. The threshold was then fixed before evaluation on a separate clinician-labeled validation set of 166 notes (91 informative, 75 non-informative), achieving 0.9578 accuracy and 0.9617 F1 (88 true-positive, 71 true-negative, 4 false-positive, and 3 false-negative notes; sensitivity 0.967, specificity 0.947, positive predictive value 0.957, and negative predictive value 0.959). This clinician-labeled validation set was not used to build the evidence library or select the threshold; because it was separate from threshold selection but not claimed here to be external by site or time, we describe it as a separate validation set rather than as an independently sourced validation set. The fixed informative-note filter was then applied to select notes for disease-activity inference; the remaining notes, which carry no disease-activity content, are not passed to the language model. Beyond this embedding filter, a second and distinct screen operates inside the agent: a note that passes the embedding filter but is found, during the agent's lightweight first-pass extraction, to still lack sufficient information for a disease-activity assessment is marked ineligible and receives no further processing. This eligibility check is intrinsic to the agent's reasoning rather than to any single backbone and is performed cheaply by the lightweight extraction model, so that costly disease-activity reasoning runs only on the notes passing both the embedding filter and this eligibility screen.

**Evidence extraction.** This stage runs as three sequential model calls — structure analysis (note sections and joint-exam template detection), objective-metric extraction, and qualitative extraction (Supplementary Methods 2). For each informative note the agent extracts the DAS28 components and related signals, recording for each not only a value but the verbatim supporting text, the note section(s) it came from, and an evidence type. It also detects whether a value was read from a structured joint-exam template (for example, a 28-joint or summary-list template) and, for swollen joints, whether the documentation specifies synovitis rather than non-specific swelling. Extracted elements include the 28-joint tender and swollen counts, the physician global assessment, C-reactive protein, any disease-activity score already documented in the note, and the patient's recent medication trajectory.

**Evidence-based integrated reasoning and quality control.** The agent assigns the primary category by one of several inference methods, each recorded explicitly: a calculated method that buckets the computed DAS28-CRP when all four components are present (remission < 2.6; low 2.6-3.2; moderate 3.2-5.1; high > 5.1); a clinical-reasoning method that applies the guideline's expert rules (for example, treating a medication escalation as evidence of higher activity) when the formula cannot be completed; and physician-assessment or conflict-resolution methods when the note documents an explicit global impression or contains reconcilable contradictions. A score-first rule is applied: when a current-visit numeric disease-activity score is documented, classification follows that score and is not down-adjusted by qualitative wording. When the evidence is conflicting or incomplete, the agent records an ambiguity flag and a secondary category, each with its own confidence and a free-text rationale. Finally, an internal validation layer runs post-inference checks and records any issues (for example, low confidence or a category inconsistent with the calculated DAS28-CRP) in an extraction-errors field, which is empty for the large majority of notes and can be used to filter lower-quality inferences.

**Supplementary Methods 2. Prompt architecture, structured outputs, and versioning**

Evidence extraction and integrated reasoning are not a single prompt. The agent is a chain of independent, individually verifiable steps; the generative steps are each constrained to emit structured JSON conforming to the schema in Supplementary Table 1. After the informative-note filter, the evidence-extraction stage is itself decomposed into three sequential prompts (structure analysis, then objective-metric extraction, then qualitative extraction); a fourth prompt performs the integrated reasoning. Decomposing the task this way is what allows a budget model to match a far more expensive model (Supplementary Methods 3, Supplementary Table 2). The sequence of calls is summarized below; representative abridged templates follow. Square brackets denote slots filled at run time; output keys are reproduced verbatim from the implementation.

| **#** | **Pipeline step** | **Purpose** | **Principal structured outputs** |
| --- | --- | --- | --- |
| 0 | Informative-note filter | Embedding retrieval over a GPT-5-derived disease-activity evidence library; non-informative notes are gated to "ineligible" before any generative call (the LLM informativeness prompt generated development labels and supporting text passages rather than serving as the final note filter). | informativeness score; is_informative; reason; supporting text |
| 1 | Structure analysis | Identify note sections (objective vs subjective) and detect/classify any 28-joint examination template. | identified_sections[]; has_28joint_template; template_type; template_text; recommended/excluded sections; confidence |
| 2 | Objective-metric extraction (structure-aware) | Extract DAS28 objective components, only from approved objective sections, with an explicit null-vs-zero rule. | tjc28_count; sjc28_count (+ sjc_synovitis_only, *_from_template); phys_global_score; crp_value; each with verbatim text, source sections, evidence_type |
| 3 | Qualitative extraction | Capture current-visit physician impression, any documented disease-activity score (by priority), and reason-based medication trajectory. | physician_impression; activity_level; existing/primary score(s); medication_status (escalated/reduced/changed/stable/unclear); medication_confidence |
| 4 | Integrated inference | Combine all evidence with the full note; apply the score-first doctrine, the DAS28-CRP calculation, and clinical-consistency conflict rules. | calculated_das28_crp; inferred_category; secondary_category (+ secondary_confidence); confidence; reasoning; inference_method |

Representative abridged template — integrated-reasoning prompt (call 4). Evidence-extraction prompts (calls 1–3) share the same structured-JSON discipline but target individual components.

SYSTEM: You are inferring a DAS28 disease-activity classification for rheumatoid arthritis from extracted clinical data and supporting evidence.
FULL CLINICAL NOTE: [note text]
EXTRACTED EVIDENCE (from calls 1-3, each with verbatim supporting text):
 TJC28=[..] SJC28=[..] Physician global=[..] CRP=[..]
 Existing DAS score=[..] Physician impression=[..]
 Medication status=[..] 28-joint template=[..]
CLASSIFICATION RULES:
 remission < 2.6; low 2.6-3.2; moderate 3.2-5.1; high > 5.1
SCORE-FIRST DOCTRINE (highest priority): if a current-visit numeric score is documented, classify strictly by it; do NOT down-adjust for wording such as 'improved' / 'stable'. Score priority: DAS28-CRP > CDAI > SDAI > DAS28-ESR > DAS28. Historical (non-current-visit) scores are invalid for classification.
CLINICAL-CONSISTENCY RULES (only when no current-visit numeric score exists): compute DAS28-CRP if all four components are present; otherwise reason from the guideline rules (e.g., treatment escalation -> higher activity). If the calculated category and the physician assessment conflict, resolve per rule and record a secondary_category with its own secondary_confidence.
OUTPUT (strict JSON): {calculated_das28_crp, initial_inferred_category, physician_assessment_category, conflict_detected, conflict_resolution_rule, reasoning, confidence_reasoning, inferred_category, confidence, secondary_category, secondary_confidence, inference_method: calculated | clinical_reasoning | physician_assessment | conflict_resolved}

**Versioning**

Each prompt, guideline revision, rule set, and model configuration is tracked in an access-controlled version-controlled repository. Every per-note output records the agent and model version used, so that any label can be traced to the exact configuration that produced it. Implementation changed iteratively during development; for reporting, adjudication-informed changes are summarized below as post hoc refinement groupings rather than as a prespecified protocol or required SHARE stopping rule. The final evaluated configuration is the version reported throughout the main text.

**Supplementary Methods 3. Comparator implementations**

**Naive single-prompt baseline.** This baseline approximates how an investigator might label notes with a general-purpose chatbot and no bespoke engineering: the full note text and the chart-review guideline are supplied together in a single prompt, and the model is asked to return the disease-activity category directly. It performs no structured evidence extraction and produces no supporting evidence, confidence, or ambiguity output, returning only a single category (or "unclear").

**Single-call agent.** This condition isolates the effect of decomposing the task into multiple steps. It uses the same task framing as SHARE-DA — structured extraction of disease-activity evidence followed by reasoning to a primary category, with a secondary category when the note is ambiguous — but performs all of it within a single model call. It therefore produces the same structured outputs as the full agent while sharing the naive baseline's single-call execution.

**Multi-step SHARE-DA agent.** The multi-step agent executes evidence extraction and integrated reasoning through four distinct, sequential model calls: structure analysis, objective-metric extraction, qualitative extraction, and integrated reasoning, so each step is simpler and individually verifiable; the final default pairs this design with a tiered backbone strategy: a lightweight model (GPT-5 Nano) performs the high-volume evidence-extraction calls, and a budget reasoning model (o4-mini, medium reasoning effort) performs the final integrated-reasoning step. Each benchmark condition applies one backbone at every step (so the GPT-5.4 conditions use GPT-5.4 for both extraction and reasoning); only the final default is tiered, pairing GPT-5 Nano extraction with the o4-mini reasoning step.

**Supplementary Table 1. SHARE-DA agent output schema**

| **Field** | **Description** |
| --- | --- |
| Note_SQL_ID | Research Patient Data Registry (RPDR) note identifier (form PatientNum__NoteID__YYYYMMDD); patient ID and note date are parsed from it. |
| Inferred DAS Category | Primary disease-activity category: remission, low, moderate, or high (or "ineligible" when the note lacks sufficient information). |
| Secondary DAS Category | Second-choice category, populated only when the note is flagged ambiguous. |
| Confidence Score / Secondary Confidence | Agent confidence (0-1) in the primary and, when applicable, the secondary category. |
| Confidence Reasoning | Free-text explanation of the confidence score. |
| TJC28 Count | Tender joint count (of 28), with the verbatim supporting text, source section(s), evidence type, and a flag for whether it was read from a structured joint-exam template. |
| SJC28 Count | Swollen joint count (of 28), with the same supporting fields plus a synovitis flag distinguishing inflammatory synovitis from non-specific swelling. |
| Physician Global Score | Physician global assessment used in DAS28-CRP, with supporting text/section and a synthesized qualitative impression. |
| CRP Value | C-reactive protein value documented in the note, with supporting text/section. |
| Existing DAS Score | A disease-activity score already documented in the note (extracted directly; not from chart review), with supporting text. |
| Calculated DAS28-CRP | DAS28-CRP computed from the formula when all four components (TJC28, SJC28, physician global, CRP) are present. |
| Inference Method | How the category was derived: calculated (from DAS28-CRP), clinical_reasoning (guideline rules, e.g., treatment escalation), physician_assessment, conflict_resolved, ineligible, and related variants. |
| Inference Reasoning | Free-text rationale for the assigned category. |
| Medication Status | Inferred RA treatment trajectory (e.g., escalated, stable) used as a clinical-reasoning signal, with supporting context and confidence. |
| Joint-exam template | Whether a 28-joint exam template was present and its type (e.g., structured_28joint, summary_list), with the template text. |
| Extraction Errors | Validation flags from the agent's internal quality-control layer (e.g., low confidence, or a category inconsistent with the calculated DAS28-CRP); empty for ~85% of notes. |
| Processing Time (s) | Wall-clock time the agent spent producing the inference for the note. |

**Supplementary Table 2. Note-level accuracy, cost, and compute time across labeling strategies and model backbones (all evaluated notes; grouped by backbone).**

| **Method** | **Backbone** | **MAE** | **Cost ($/note)** | **Time (s/note)** |
| --- | --- | --- | --- | --- |
| Multi-step agent (final, default) | o4-mini-medium | 0.291 | 0.0670 | 82.7 |
| Multi-step agent (initial) | o4-mini-medium | 0.406 | 0.0670 | 82.7 |
| Single-call agent | o4-mini-medium | 0.349 | 0.00998 | 14.5 |
| Naive (single-prompt) | o4-mini-medium | 0.345 | 0.0055 | 9.1 |
| Multi-step agent | GPT-5.4-high | 0.310 | 0.2193 | 275.6 |
| Single-call agent | GPT-5.4-high | 0.332 | 0.0462 | 57.7 |
| Naive (single-prompt) | GPT-5.4-high | 0.341 | 0.0149 | 13.4 |

*MAE = primary+secondary blended, vs the final co-produced labels (remission=1...high=4). Rows are labeled by their reasoning backbone, which determines the assigned category and hence accuracy: the GPT-5.4 conditions apply GPT-5.4 at every step, whereas the final default additionally runs the high-volume extraction step on GPT-5 Nano—lowering cost and time without changing the o4-mini reasoning step. The final (0.291) and initial (0.406) agent rows therefore share the same o4-mini reasoning backbone and differ only by co-learning.*

Benchmark results. Across all evaluated notes, the final multi-step agent on the budget o4-mini backbone achieved the lowest MAE of any configuration tested (0.291) — being comparable, or even slightly better than the same multi-step agent run on the far more capable GPT-5.4 at high reasoning effort (0.310), at under a third of the per-note cost and under a third of the compute time. This budget-tier accuracy was observed after adjudication-informed refinements: with the backbone held fixed, the o4-mini agent changed from 0.406 to 0.291 between the initial and final evaluated configurations (Supplementary Table 2). The advantage came from how the task was decomposed rather than from raw model power. Multi-step labeling was the most accurate configuration for both backbones, and on the capable GPT-5.4 the expected ordering held throughout (multi-step 0.310 < single-call 0.332 < naive single-prompt 0.341); but on the weak o4-mini only full multi-step decomposition was effective — the single-call agent (0.349) did no better than the naive prompt (0.345), and both trailed far behind multi-step (0.291). Decomposition thus helped the weak backbone far more than the strong one (an MAE reduction of roughly 0.05 versus 0.02–0.03, from Supplementary Table 2), consistent with a capable model already reasoning well in a single call: breaking the task into smaller, individually verifiable steps substitutes for model strength, letting a cheap backbone match an expensive one — exactly what budget-constrained labeling at scale requires.

During adjudication, recurrent discrepancy patterns were used to refine the agent prompts and abstraction guidance across three adjudication rounds. These rounds describe the sequence of prompt refinement during development rather than a prespecified SHARE protocol or stopping rule. In Round 1, prompt revisions strengthened joint-exam interpretation: the agent was instructed to recognize structured 28-joint, anatomical-list, summary-list, and abbreviated joint-group templates; distinguish a true joint-exam template from a bare section header; identify the section source of template content; and apply DAS28 joint inclusion and exclusion rules when templates mixed DAS28 and non-DAS28 joints. In Round 2, prompt revisions addressed evidence integration and conflict resolution: the agent was instructed to prioritize current-visit numeric disease-activity scores when available, distinguish current from historical scores and treatment decisions, resolve conflicts between calculated scores and physician assessments using prespecified clinical rules, and return a secondary category when an alternative interpretation remained plausible. In Round 3, prompt revisions targeted recurrent special cases observed during adjudication, including clinician-documented objective pain or discomfort versus patient-reported symptoms alone, squeeze tenderness as evidence of tenderness, fullness or synovitis as evidence of swelling, non-RA or non-DAS28 findings that should not drive the RA disease-activity category, and medication escalation or continuation only when tied to the current visit. These prompt-level refinements defined the final evaluated SHARE-DA configuration used for large-scale inference.

Agent note-level accuracy before and after co-learning is reported in main-text Table 2. The MAE across all notes is not reported for the initial human labels. The co-produced reference was created by revising those labels note by note, so on the ~80% of notes that adjudication left unchanged the initial label equals the reference by construction; a note-level MAE is therefore driven mechanically toward zero and would overstate the accuracy of the initial human read. The initial human labels are compared only where the contrast is informative — on the adjudicated notes and at the visit (window) level (Supplementary Table 3).

**Supplementary Table 3. Window-level accuracy against expert review**

Note: Data from Table below presented in Figure 2. Data duplicated here to more clearly link the results with the corresponding methods.

| **Approach** | **MAE (vs co-produced reference)** |
| --- | --- |
| Human initial review | 0.280 |
| Initial agent | 0.424 |
| Final agent | 0.281 |

414 recency-weighted patient–timepoint windows at two timepoints (the b/tsDMARD switch and one year afterward); within each window, notes are weighted by recency to the timepoint. Reported at the visit level — how disease activity is used in downstream studies — alongside the note-level analysis, for studies needing per-visit summaries.

This visit-level comparison aggregates each method into its own recency-weighted per-visit summary rather than matching labels note by note, so it does not inherit the note-level circularity in which the initial labels and the reference coincide on the unrevised majority of notes; the resulting initial-human window MAE (0.280) reflects genuine visit-level divergence from the reference, making the final agent's match to it (0.281 vs 0.280) a fair comparison.

The final agent's weighted MAE against the co-produced reference was 0.45 on the notes reviewers flagged ambiguous versus 0.28 on the non-ambiguous notes (overall note-level MAE 0.291; Table 2).
